# Comparative analysis of Respiratory Syncytial Virus frequency rates and viral load in different patient cohorts in a university hospital in Sao Paulo, Brazil, over an eight-year period (2005-2013)

**DOI:** 10.1101/2023.06.29.23292044

**Authors:** Luciano Kleber de Souza Luna, Jessica Santiago Cruz, Tânia do Socorro Souza Chaves, Nancy Bellei

## Abstract

Respiratory Syncytial Virus (RSV) poses a global health concern, particularly in young children, the elderly, and those immunosuppressed. RSV viral load is essential for understanding transmission, disease severity, prevention, and treatment. This retrospective study aimed to analyze the frequency rates and viral loads of RSV infections in different patient cohorts and age groups over an eight-year period in a university hospital in São Paulo, Brazil. This study analyzed 1380 immunocompetent (IC) and immunosuppressed (IS) patients with acute respiratory tract infections. IC included patients suspected of having severe acute respiratory syndrome caused by influenza A (H1N1)pdm09 virus (SARS H1N1), chronic heart disease (HD), and those receiving primary care service (PC). The IS comprised transplant patients and individuals with human immunodeficiency virus (HIV) infection. Respiratory samples were collected between 2005 and 2013. RSV detection and quantification were performed by RT-qPCR. Viral load was expressed as Log10 copies of RNA/mL. The overall RSV infection rate was 17.3%, with higher rates in children (23.9%) than in adults (12.9%), particularly in children under two years of age (28.2%). Children in the SARS H1N1 and PC subgroups had higher infection rates (16.4% and 34.9%, respectively), with the highest rate in PC children aged 1 to <2 years (45.45%). Adults with HD had a significantly higher frequency rate (27.83%) than those in the SARS H1N1 (2.65%) and IS (15.16%) subgroups. The RSV viral load ranged from 2.43 to 10.15 Log10 RNA copies/mL (mean ±SD = 5.82 ± 2.19). Hospitalized patients had significantly higher viral loads (7.34 ± 1.9) than outpatients (4.38 ± 1.89). Elderly bone marrow transplant patients also had significantly higher viral loads (7.57 ± 2.41) than younger adults (5.12 ± 1.87). This study provides insights into the patterns and impacts of RSV infection in different patient cohorts in Brazil, contributing to developing effective prevention, treatment, and control strategies. Further investigations are needed to understand the susceptibility and risk factors associated with RSV infection.

## Introduction

Respiratory Syncytial Virus (RSV) poses a significant global health concern due to its high prevalence and the potential for severe disease outcomes, particularly in vulnerable populations such as young children, elderly individuals, and those with compromised immune systems ^1–5^.

Analyzing the RSV viral load is crucial for gaining insight into the clinical course, disease severity, and transmission dynamics of RSV infections. Viral load measurement provides valuable information regarding the replication kinetics of the virus, shedding patterns, and host immune responses, all of which can significantly affect disease outcomes and transmission potential ^6,7^. Assessment of the viral load can provide valuable insights into the patterns and impact of RSV infection, contributing to the establishment of effective strategies for prevention, treatment, and control. Current treatment options are supportive; however, antiviral trials are ongoing and new preventive options are available. Studies on the development of monoclonal antibodies against RSV have also advanced. Niservimab has emerged as an additional option to prevent RSV infections in preterm and term infants ^8–10^. In addition, The US Food and Drug Administration (FDA) recently approved an RSV vaccine for use in the United States in individuals aged 60 years and older ^11^. Understanding the natural dynamics of viral infection and replication is crucial before new therapeutic interventions become available in the market.

The viral load of RSV can vary substantially among infected individuals and is influenced by multiple factors such as age, immune status, and comorbidities. Among young children who are particularly susceptible to severe RSV infections, higher viral loads have been associated with more severe clinical manifestations, including bronchiolitis and pneumonia ^1,6,12–15^. These findings underscore the importance of viral load as a determinant of disease severity, and highlight the need for early identification and intervention in high-risk pediatric populations.

Elderly individuals, especially those with underlying health conditions, are at increased risk of developing severe RSV disease. Studies have shown that the viral loads in elderly adults with RSV infections are often higher than those in younger adults, which may contribute to the greater disease severity observed in this age group ^1,16^.

Immunocompromised individuals, such as those undergoing transplantation or living with human immunodeficiency virus (HIV) infection, are particularly vulnerable to severe RSV infections ^3^. This emphasizes the significance of viral load as a potential parameter for assessing disease risk and developing targeted interventions for the elderly population.

This retrospective study aimed to analyze the frequency rates and viral load of RSV infections in a diverse cohort of patients with acute respiratory tract infections.

## Material and Methods

### Patients

A total of 1380 patients who sought medical care for acute respiratory tract infections (ARI) were retrospectively analyzed. Patients from different cohorts were treated at São Paulo University Hospital, a tertiary hospital in the city of São Paulo, Brazil, from 2005 to 2013. Clinical and demographic data were collected from the medical records to characterize the patients. The cohorts were divided into two main groups: immunocompetent or IC (n=924) and immunosuppressed or IS (n=456).

Within the IC group, further subgroup analysis was conducted to evaluate the viral load in specific patient populations, including (i) 520 individuals suspected of having severe acute respiratory syndrome caused by influenza A (H1N1)pdm09 virus (SARS H1N1); (ii) 118 children with chronic heart disease (cHD); (iii) 101 adults with chronic heart disease (aHD); and (iv) 185 individuals who received primary care service (PC). Patients aged less than 13 years were considered children. The IS group consisted of 355 transplant patients, including 282 bone marrow (BMT), 44 kidney (KT), 10 liver (LT), and 120 individuals with human immunodeficiency virus (HIV) infection.

SARS H1N1 and PC patients were also divided into the following age groups: <0,5; 0,5 to <1, 1 to <2, 2 to <5, and 5 to <13.

The sample collection period for each group is shown in Fig. 1.

**Fig. 1.**
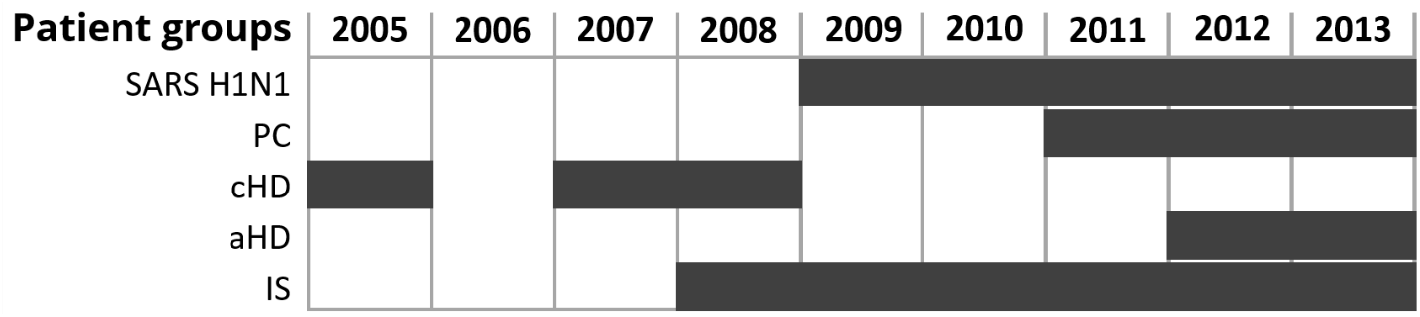
Patient groups and sample collection period. SARS H1N1, suspected of having severe acute respiratory syndrome caused by influenza A (H1N1)pdm09 virus. PC, primary care. cHD, children with heart disease. aHD, adults with heart disease. IS, immunosuppressed.

### Sample collection

Nasopharyngeal swabs were collected from all patients, except for children under two years of age, for whom samples were obtained from nasopharyngeal aspirates. The collected specimens were placed in 2 mL of sterile lactated Ringer’s solution, homogenized, and aliquoted before storage at - 80°C.,

### RNA purification

RNA was purified from aliquots of respiratory samples stored at -80°C using the QIAamp Viral RNA Mini Kit (Qiagen, Hilden, Germany), according to the manufacturer’s instructions.

### Respiratory Syncytial Virus detection and quantification

RSV detection and quantification were achieved through a one-step quantitative reverse transcription-polymerase chain reaction (RT-qPCR) with oligonucleotides targeting a conserved region of the RSV matrix gene ^17^.

The human ribonuclease P (RNase P) gene was amplified in a parallel reaction to assess RNA purification efficacy and as an internal control using oligonucleotides described elsewhere ^18^. RT-qPCRs were performed in a total volume of 25 μL using AgPath-ID One-Step RT-PCR Reagents (Applied Biosystems, Austin, USA) with 5 μL of purified RNA. The RSV RT-qPCR contained 900 nM of each primer and 200 nM of the TaqMan probe. For RNase P RT-qPCR, 800 nM of each primer and 200 nM of TaqMan probe were used. The reactions were performed on an Applied Biosystems 7500 Real-Time PCR System (Applied Biosystems) for 10 min at 95°C and 10 min at 50°C, followed by 45 cycles of 10 s at 95°C, and 30 s at 55°C (data collection). The cycle threshold cut-off value for RSV-positive detection was established as ≤ 40.

The viral load or RSV was determined by interpolating the cycle threshold values into a standard curve generated from triplicate 10-fold serial dilutions of the quantified *in vitro transcribed* RNA containing the amplification target. Viral load values were expressed as Log10 copies of RNA/mL.

### Ethics

This study was approved by the Research Ethics Committee of São Paulo Hospital and Universidade Federal de São Paulo (CEP/UNIFESP n. 0086/2016, CAAE n. 53012516.8.0000.5505).

### Statistics

Continuous variables were summarized as mean ± standard deviation, interquartile range (IQR: Q3-Q1), and median. Categorical variables were described as frequency rates. As appropriate, statistical analyses were performed using Fisher’s exact test and the chi-square, Mann-Whitney, Kruskal-Wallis, and Dunn’s multiple comparison tests. Statistical significance was set at *P* < 0.05. All analyses were conducted using GraphPad Prism v.6.01 (GraphPad Software, California, USA).

## Results

### RSV detection

The study included patients aged 0-91 years, with a mean age of 28.1 ± 24.9 (IQR:49–1) and a median age of 27. Age distribution by subgroup is shown in Table 1.

**Table 1.** Age distribution by subgroup of patients

| Patient subgroup | Number | Age (years old) |  |  |
| --- | --- | --- | --- | --- |
| | | Range | Mean ± SD (IQR) | Median |
| SARS H1N1 | 520 | 0 - 91 | 23.6 ± 25.3 (42 - 1) | 13 |
| <0,5 | 47 | 0 - 0.42 | 0.23 ± 0.13 (0.33 - 0.08) | 0.25 |
| 0,5 - <1 | 46 | 0.5 - 0.92 | 0.68 ± 0.14 (0.81 - 0.56) | 0.67 |
| 1 - <2 | 48 | 1 - 1.83 | 1.03 ± 0.16 (1 - 1) | 1 |
| 2 - <5 | 65 | 2 - 4 | 2.63 ± 0.74 (3 - 2) | 2 |
| 5 - <13 | 50 | 5 - 12 | 7.7 ± 2.15 (9.25 - 6) | 7 |
| 13 - <65 | 215 | 13 - 64 | 36.97 ± 14.37 (51 - 24) | 36 |
| $\geq 65$ | 49 | 65 - 91 | 75.24 ± 7.65 (82 - 68.5) | 74 |
| PC | 185 | 0 - 86 | 4.3 ± 11.8 (1 - 0.2) | 0.58 |
| <0,5 | 80 | 0 - 0.33 | 0.2 ± 0.13 (0.25 - 0.08) | 0.17 |
| 0,5 - <1 | 39 | 0.5 - 0.92 | 0.67 ± 0.13 (0.75 - 0.58) | 0.67 |
| 1 - <2 | 22 | 1 - 1 | 1 ± 0.0 (1 - 1) | 1 |
| 2 - <5 | 13 | 2 - 4 | 2.46 ± 0.78 (3 - 2) | 2 |
| 5 - <13 | 15 | 5 - 12 | 7.6 ± 2.82 (11 - 5) | 7 |
| 13 - <65 | 14 | 14 - 54 | 29.93 ± 11.58 (38.5 - 20.5) | 28.5 |
| $\geq 65$ | 2 | 79 - 86 | 82.5 ± 4.95 (86 - 79) | 82.5 |
| aHD | 101 | 13 - 79 | 56.6 ± 13.3 (66.5 - 48) | 58 |
| cHD | 118 | 0 - 11 | 2.9 ± 3.04 (5 - 0.7) | 1.29 |
| IS | 456 | 0 - 84 | 43 ± 15.3 (53 - 33) | 44 |
| IS-BMT | 282 | 17 - 84 | 43 ± 14.7 (43 - 31) | 42 |
| IS-KT | 44 | 0 - 80 | 47.9 ± 16.2 (58 - 38.3) | 50 |
| IS-LT | 10 | 0 - 56 | 7.67 ± 17.2 (4.5 - 0.75) | 1.5 |
| IS-HIV | 120 | 0 - 71 | 44.1 ± 12.4 (52 - 38) | 44.5 |

SARS H1N1, suspected of having severe acute respiratory syndrome caused by influenza A (H1N1)pdm09 virus. PC, primary care. aHD, adults with heart disease. cHD, children with heart disease. IS, immunosuppressed. BMT, bone marrow transplant. KT, kidney transplant. LT, liver transplant. HIV, human immunodeficiency virus-positive. SD, standard deviation. IQR, interquartile range.

The IS cohort comprised mostly adults (95.8%), whereas the PC cohort predominantly comprised children (91.3%).

In total, 239 of the 1380 patients (557 children and 823 adults) tested positive for RSV (17.3%). The frequency rates in the cohorts analyzed by age group are shown in Table 2.

**Table 2.** RSV frequency rates according to subgroups and age groups.

| Subgroups by age (years) | Number | RSV |  |  |
| --- | --- | --- | --- | --- |
|  |  | Positive | Negative | Frequency rate (%) |
| SARS H1N1 | 520 | 49 | 471 | 9.42 |
| <0,5 | 47 | 10 | 37 | 21.28 |
| 0,5 - <1 | 46 | 13 | 33 | 28.26 |
| 1 - <2 | 48 | 9 | 39 | 18.75 |
| 2 - <5 | 65 | 8 | 57 | 13.31 |
| 5 - <13 | 50 | 9 | 48 | 4 |
| 13 - <65 | 215 | 6 | 209 | 2.79 |
| ≥ 65 | 49 | 1 | 48 | 2.04 |
| PC | 185 | 61 | 124 | 32.97 |
| <0,5 | 80 | 30 | 50 | 37.5 |
| 0,5 - <1 | 39 | 15 | 24 | 38.46 |
| 1 - <2 | 22 | 10 | 12 | 45.45 |
| 2 - <5 | 13 | 3 | 10 | 23.08 |
| 5 - <13 | 15 | 1 | 14 | 6.67 |
| 13 - <65 | 14 | 1 | 13 | 7.14 |
| ≥ 65 | 2 | 1 | 1 | 50 |
| cHD | 118 | 30 | 88 | 25.4 |
| aHD | 101 | 30 | 71 | 29.7 |
| 13 - <65 | 68 | 20 | 48 | 29.41 |
| ≥ 65 | 29 | 7 | 22 | 24.13 |
| IS | 456 | 69 | 387 | 15.1 |
| 0 < 13 | 14 | 2 | 12 | 14.29 |
| 13 - <65 | 407 | 60 | 347 | 14.74 |
| ≥ 65 | 35 | 7 | 28 | 20 |
| IS-BMT | 282 | 53 | 229 | 18.79 |
| IS-KT | 44 | 2 | 42 | 4.55 |
| IS-LT | 10 | 1 | 9 | 10 |
| IS-HIV | 120 | 13 | 107 | 10.83 |

SARS H1N1, suspected of having severe acute respiratory syndrome caused by influenza A (H1N1)pdm09 virus. PC, primary care. aHD, adults with heart disease. cHD, children with heart disease. IS, immunosuppressed. BMT, bone marrow transplant. KT, kidney transplant. LT, liver transplant. HIV, human immunodeficiency virus-positive. SD, standard deviation. IQR, interquartile range.

The difference between the RSV infection rates of immunocompetent (18.4%) and immunosuppressed (15.1 %) patients was not significant (*p* = 0.1506).

The RSV frequency was significantly higher (*p* < 0.0001) in children (23.9%) than in adults (12.9%). In general, children in aged groups under two years of age exhibited significantly (*p* = 0.0001) higher infection rates than older children (aged 5 to < 13 years).

Subgroup analysis revealed variations in RSV infection rates among children aged 0 to 5 years old in SARS H1N1 patients. In this subgroup, children aged from 0.5 to <1 year presented the highest frequency rate (28.26%), which was significantly higher (*p*=0.048) than that in children aged 2 to <5 years (13.31%).

Among SARS H1N1 and PC, children aged 5 to < 13 years had the lowest frequency rates compared to the other children age groups (4% and 6.67%, respectively), whereas PC children aged 1 - <2 years showed the highest frequency rate (45.45%). This frequency of infection was significantly higher than SARS H1N1 children aged < 0.5 (*p*=0.049), 1 to < 2 years (*p*=0.040), and 2 to < 5 years (*p*=0.002).

Children in the SARS H1N1 and PC subgroups were significantly more likely to be infected than adults (odds ratio = 7.2 and 3.75, respectively). The frequency rates in the analyzed cohorts of children and adults are shown in Table 3.

**Table 3.**
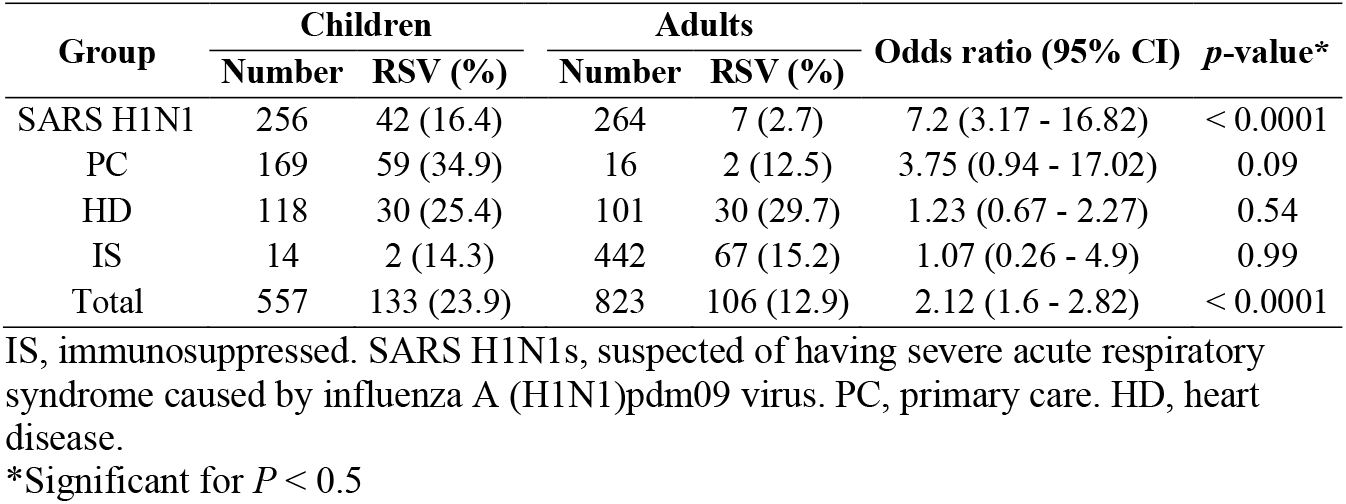
RSV frequency rates and odds ratios of infection in children and adults.

| Group | Children |  | Adults |  | Odds ratio (95% CI) | p-value* |
| --- | --- | --- | --- | --- | --- | --- |
|  | Number | RSV (%) | Number | RSV (%) |  |  |
| SARS H1N1 | 256 | 42 (16.4) | 264 | 7 (2.7) | 7.2 (3.17 - 16.82) | < 0.0001 |
| PC | 169 | 59 (34.9) | 16 | 2 (12.5) | 3.75 (0.94 - 17.02) | 0.09 |
| HD | 118 | 30 (25.4) | 101 | 30 (29.7) | 1.23 (0.67 - 2.27) | 0.54 |
| IS | 14 | 2 (14.3) | 442 | 67 (15.2) | 1.07 (0.26 - 4.9) | 0.99 |
| Total | 557 | 133 (23.9) | 823 | 106 (12.9) | 2.12 (1.6 - 2.82) | < 0.0001 |

IS, immunosuppressed. SARS H1N1s, suspected of having severe acute respiratory syndrome caused by influenza A (H1N1)pdm09 virus. PC, primary care. HD, heart disease.
\*Significant for $P < 0.5$

Adults with HD presented the highest frequency rate (27.83%), which was significantly higher (*p*<0.0001) than those in the SARS H1N1 (2.65%) and IS (15.16%, *p*=0.0047) subgroups but not in comparison to those in the PC subgroup (12.5%, *p*=0.234).

In the IS group, RSV infection rates varied across different transplant patients, with BMT patients showing the highest infection rate of 18.8% (53/282), followed by 4.5% in KT patients (2/44), and 10% in LT patients (1/10). Among patients with HIV infection, 10.8% were positive (13/120).

Most IS patients were hospitalized (63.3%), but no RSV significant difference (*p* = 0.507) between hospitalized (17.8%) and outpatients (16.4 %).

There were no significant differences in RSV frequency rates between hospitalized and outpatient children. In adults, the frequency rates were significantly higher in outpatients with heart disease (*p*<0.0001) and in immunosuppressed hospitalized patients (p=0.032).

### RSV Viral Load

The viral load of RSV-infected samples was calculated by interpolating their *Ct* values into the equation obtained from the RSV RT-qPCR standard curve (Fig. 2), and was expressed as Log10 RNA copies/mL.

**Fig. 2.**
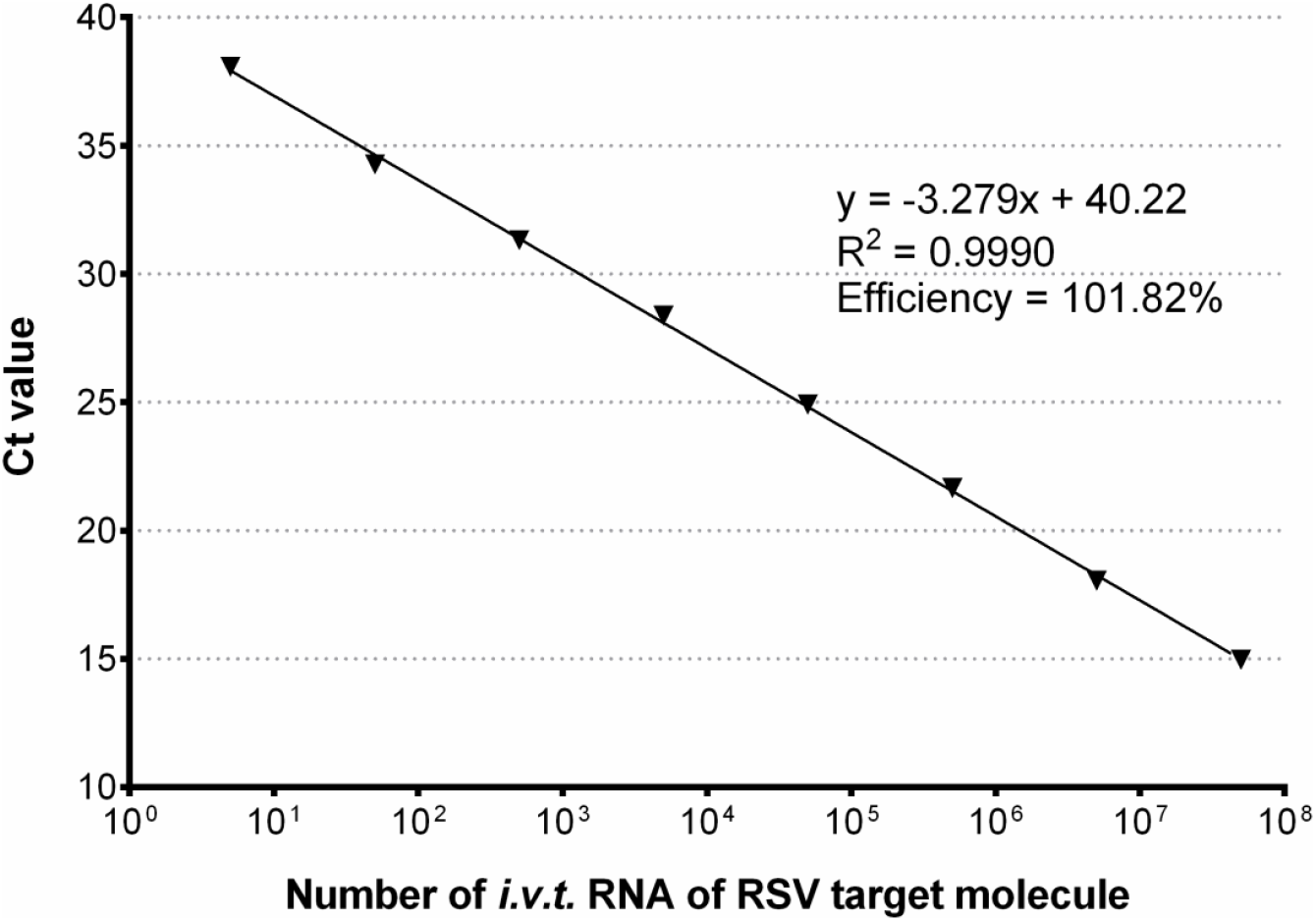
Generated standard curve to determine the viral load of RSV using serial dilutions of quantified *in vitro transcribed (i*.*v*.*t)*. RNA containing the RSV RT-qPCR amplification target. The value for each dilution is the average ± standard deviation of three replicates.

The overall viral load of RSV infected patients ranged from 2.43 to 10.15 Log10 RNA copies/mL, with a mean of 5.82 ± 2.19 (IQR: 7.63-3.53) and a median of 5.85. Viral load distribution by subgroup is shown in Table 4.

**Table 4.** Viral load distribution by patient subgroup and age group

| Patient subgroup | Number | Viral load (Log10 RNA copies/mL) |  |  |
| --- | --- | --- | --- | --- |
| | | Range | Mean ± SD (IQR) | Median |
| SARS H1N1 | 49 | 3.63 - 9.16 | 6.94 ± 1.36 (7.93 - 5.98) | 7.30 |
| <0,5 | 10 | 4.71 - 8.53 | 7.00 ± 1.25 (7.78 - 6.12) | 7.42 |
| 0,5 - <1 | 13 | 3.63 - 8.36 | 6.71 ± 1.41 (7.75 - 5.67) | 6.98 |
| 1 - <2 | 9 | 5.85 - 8.98 | 7.75 ± 1.01 (8.60 - 7.09) | 7.95 |
| 2 - <5 | 8 | 4.84 - 8.25 | 6.87 ± 1.23 (7.85 - 5.66) | 7.29 |
| 5 - <13 | 2 | 4.88 - 8.76 | 6.82 ± 2.74 (8.76 - 4.88) | 6.82 |
| 13 - <65 | 6 | 5.37 - 9.16 | 6.66 ± 1.39 (7.81 - 5.64) | 6.19 |
| $\geq 65$ | 1 | 4.25 | - | 4.25 |
| PC | 61 | 3.74 - 10.0 | 7.32 ± 1.70 (8.87 - 6.18) | 7.39 |
| <0,5 | 30 | 4.18 - 9.62 | 7.83 ± 1.48 (9.02 - 6.85) | 8.81 |
| 0,5 - <1 | 15 | 3.74 - 10.04 | 7.37 ± 1.77 (8.98 - 6.53) | 7.16 |
| 1 - <2 | 10 | 4.45 - 9.09 | 6.89 ± 1.69 (8.22 - 5.12) | 7.42 |
| 2 - <5 | 3 | 4.28 - 6.75 | 5.79 ± 1.33 (6.75 - 4.28) | 6.35 |
| 5 - <13 | 1 | 6.14 | - | 6.14 |
| 13 - <65 | 1 | 3.89 | - | 3.89 |
| $\geq 65$ | 1 | 4.61 | - | 4.61 |
| aHD | 30 | 2.63 - 5.47 | 3.35 ± 0.51 (3.48 - 3.05) | 3.28 |
| cHD | 30 | 2.43 - 9.63 | 4.26 ± 1.97 (4.79 - 3.03) | 3.37 |
| IS | 69 | 2.73 - 10.2 | 5.46 ± 2.09 (7.28 - 3.52) | 7.12 |
| IS-BMT | 53 | 3.12 - 10.5 | 5.62 ± 2.05 (7.45 - 3.69) | 5.16 |
| IS-KT | 2 | 7.27 - 8.06 | 7.67 ± 0.56 (8.06 - 7.27) | 7.67 |
| IS-LT | 1 | 9.25 | - | - |
| IS-HIV | 13 | 2.73 - 7.62 | 4.16 ± 1.59 (4.77 - 3.17) | 3.51 |

SARS H1N1, suspected of having severe acute respiratory syndrome caused by influenza A (H1N1)pdm09 virus. PC, primary care. aHD, adults with heart disease. cHD, children with heart disease. IS, immunosuppressed. BMT, bone marrow transplant. KT, kidney transplant. LT, liver transplant. HIV, human immunodeficiency virus-positive. SD, standard deviation. IQR, interquartile range.

The mean RSV viral loads in the PC and SARS H1N1 subgroups were significantly higher than those in other subgroups. Hospitalized patients had significantly higher (*p*<0.001) RSV viral loads (7.34 ± 1.9, IQR: 8.97 - 6.16) compared to outpatients (4.38 ± 1.89, IQR: 5.74 - 3.06). The same pattern was observed (*p*=0.047) for hospitalized (4.55 ± 1.82, IQR: 5.47 - 3.12) and outpatient adults (3.54 ± 0.86, IQR: 3.54 - 3.02).

Within the subgroups, only IS hospitalized patients presented significantly higher (*p*=0.0002) mean RSV viral loads (6.03 ± 2.1, IQR: 7.69 - 4.09) than IS outpatients (4.04 ± 1.23, IQR: 4.19 - 3.45) (Fig. 3).

**Fig. 3.**
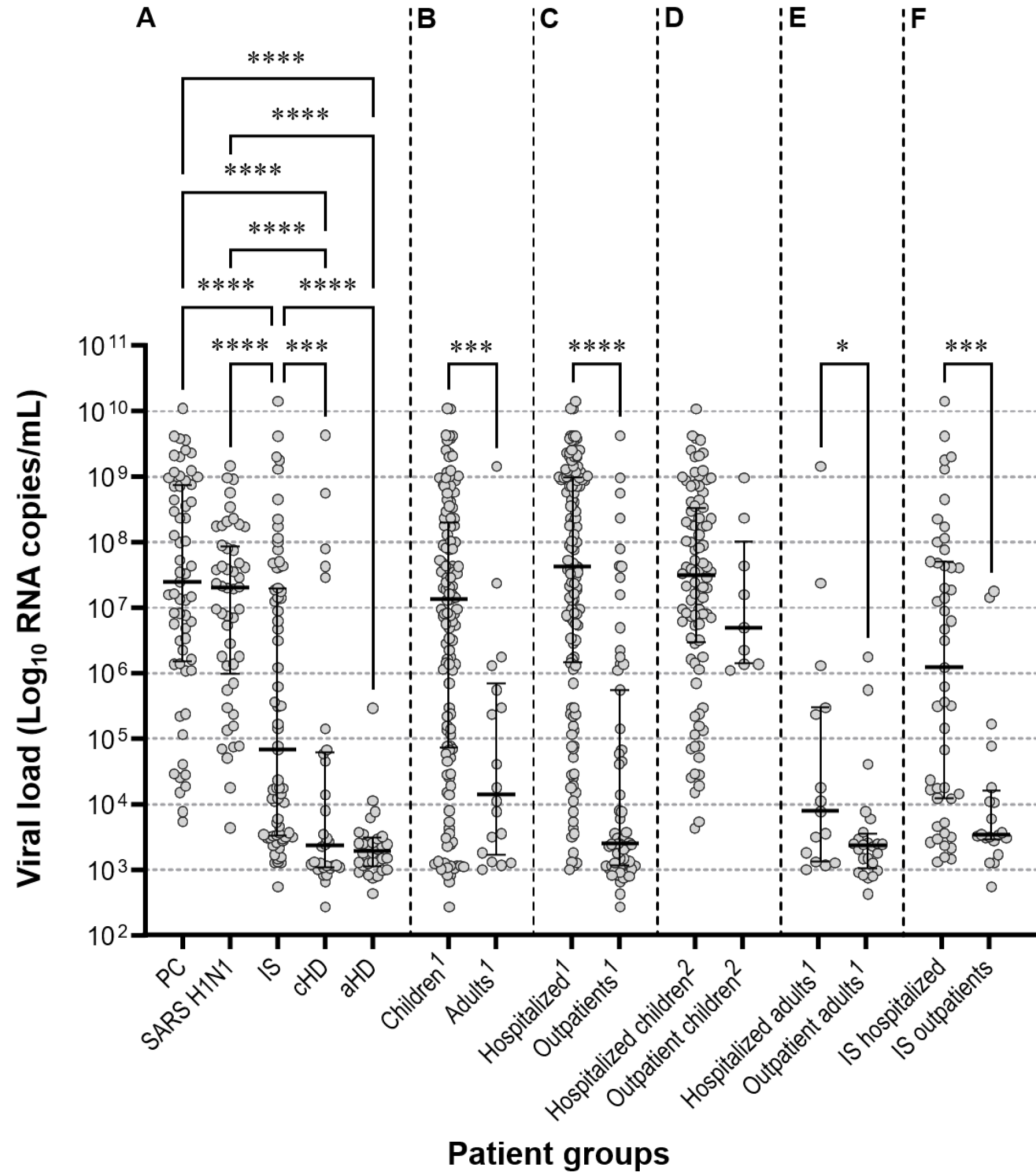
Scatter plots with median and interquartile ranges showing the RSV viral load distribution. A, studied groups. B, children and adults. C, hospitalized patients and outpatients. D, hospitalized and outpatient children. E, hospitalized and outpatient adults. F, hospitalized and immunosuppressed outpatients. PC, primary care. SARS H1N1, suspected of having severe acute respiratory syndrome caused by influenza A (H1N1)pdm09 virus. IS, immunosuppressed. aHD, adults with heart disease. cHD, children with heart disease. ^1^ Immunosuppressed not included. ^2^ children with heart disease not included.

There were no significant differences in RSV mean viral loads within SARS H1N1 and PC children in all children age groups or among these subgroups.

A comparison between younger adults (aged 18-64 years) and the elderly (≥ 65 years) was only possible in bone marrow transplant patients due to the absence or small number of elderly individuals in the other subgroups. Younger adults had a mean RSV viral load of 5.12 ± 1.87 (IQR: 7.05-3.5), whereas the elderly had a significantly higher (*p*=0.0079) mean RSV viral load of 7.57 ± 2.41 (IQR: 9.62-5.16).

Children and adults with heart disease exhibited significantly lower mean RSV viral loads than those in the SARS H1N1, PC, and IS subgroups (figure 4).

## Discussion

This retrospective study evaluated RSV frequency rates and viral loads in a diverse group of patients with acute respiratory tract infections over an eight-year period. The study population comprised both immunocompetent and immunosuppressed individuals, including those with different age ranges and chronic heart disease. The immunosuppressed (IS) subgroup was almost entirely composed of adults (95.8%), whereas the primary care (PC) subgroup was almost entirely comprised of children (91.3%), thereby hindering comparisons between children and adults within these subgroups.

The overall RSV infection rate was 17.3%, with no significant difference between the immunocompetent and immunosuppressed patients. However, the RSV frequency was significantly higher in children than in adults, particularly in children under two years of age, compared to older children. Additionally, subgroup analysis revealed varying infection rates among the different patient cohorts, with higher rates observed in children in the SARS H1N1 and PC subgroups. Notably, children in the PC subgroup aged 1 to < 2 years presented the highest RSV frequency rate, whereas children aged 5 to < 13 years, in both SARS H1N1 and PC subgroups, had the lowest frequency rates compared to younger children. These findings are consistent with previous studies that have reported higher RSV infection rates in infants compared to children and adults ^19,20^, and suggest that certain subpopulations within the immunocompetent group may be more susceptible to RSV infections owing to increased exposure to RSV in community settings, requiring further investigation and strategic interventions ^19^.

The mean RSV viral load was higher in the PC and SARS H1N1 subgroups compared to other subgroups. Similarly, children had a higher mean RSV viral load than adults. Hospitalized patients, both children and adults, exhibited significantly higher RSV viral loads than outpatients, and experienced more severe disease manifestations, as evidenced by the higher hospitalization rates. These results are consistent with those of previous studies that demonstrated comparable results ^5,12,16^.

Among immunosuppressed patients, hospitalized individuals had significantly higher mean RSV viral loads than outpatients. Notably, bone marrow transplant patients in the elderly age group had a significantly higher mean RSV viral load than younger adults, suggesting that these populations are more susceptible to severe RSV infections ^20^. This outcome may be explained by age-related changes in the immune system (immune senescence), and impaired antiviral defense mechanisms may contribute to the higher viral loads and greater disease severity observed in older individuals ^20–22^.

Patients in the heart disease subgroup (HD) had significantly lower mean RSV viral loads than those in the other subgroups, although children on HD had the second highest RSV frequency rate among children, and adults on HD had the highest among adults. In children with RSV-acute lower respiratory infections, congenital heart disease is a risk factor for a severe clinical course, but a reduced RSV viral load may be associated with a lower immune response and prolonged virus clearing, leading to worse outcomes ^23,24^. However, further research is needed to unravel the complex interplay among comorbidities, viral load, and disease outcomes in RSV-infected individuals.

This study had some limitations. This study was conducted at a single center, which may restrict the generalizability of the results to other populations or regions, highlighting the need for caution when extrapolating these findings to different healthcare settings or populations with distinct characteristics.

Additionally, the retrospective nature of the study introduces inherent limitations, such as incomplete or missing information in medical records, potentially impacting the accuracy and completeness of the findings. The availability and consistency of the data across the study period may have varied, thereby influencing the analysis and interpretation of the results.

Furthermore, the relatively small sample size, particularly in specific subgroups, such as patients with specific comorbidities or transplant recipients, may limit the ability to detect significant differences in viral load between these subgroups and restrict the generalizability of the findings to larger populations.

Another limitation is that this study focused solely on RSV viral load and its association with clinical outcomes, neglecting other factors influencing disease severity and transmission dynamics, such as the host immune response, viral genetic variability, and environmental factors. Future studies should consider a more comprehensive approach to provide a broader understanding of the RSV infection dynamics.

In conclusion, despite these limitations, the findings of this study provide valuable insights into the role of viral load as a determinant of disease severity in different age groups and immune statuses in Brazil. Understanding the dynamics of viral load in specific subpopulations can contribute to the development of targeted interventions to improve the prevention, treatment, and control strategies for RSV infections.

## Data Availability

All data produced in the present work are contained in the manuscript

